# FOCUS: Forecasting COVID-19 in the United States

**DOI:** 10.1101/2021.05.18.21257386

**Authors:** VP Nagraj, Chris Hulme-Lowe, Stephanie L. Guertin, Stephen D. Turner

## Abstract

Infectious disease forecasting has been a useful tool for public health planning and messaging during the COVID-19 pandemic. In partnership with the CDC, the organizers of the COVID-19 Forecast Hub have created a mechanism for forecasters from academia, industry, and government organizations to submit weekly near-term predictions of COVID-19 targets in the United States. Here we describe our efforts to participate in the COVID-19 Forecast Hub through the **Fo**recasting **CO**VID-19 in the **U**nited **S**tates (**FOCUS**) project. The effort led to more than three months of weekly submissions and development of an automated pipeline to generate forecasts. The models used in FOCUS yielded forecasts that ranked relatively well in terms of precision and accuracy.

## 1 Background

The COVID-19 pandemic has demonstrated that rigorous statistical assessment of disease dynamics can and should help inform public health efforts. In order to “flatten the curve,” public health authorities need to anticipate the epidemic trajectory over time. Infectious disease forecasting can help guide understanding of the effect of population-wide interventions and activities such as statewide lock downs, commercial re-openings, or even differential allocation of healthcare resources. The field of epidemiological modeling is mature, and the methods developed have been critical in responding to outbreaks of many different diseases including malaria, dengue fever, Zika, AIDS, and Ebola. In recent years, there has been a great deal of work focused on seasonal influenza, which yearly kills thousands and sickens millions more around the world. The Centers for Disease Control (CDC) has coordinated national, regional, and statewide influenza surveillance efforts in the United States for years. Since 2013, the CDC has invited experts in epidemiology, influenza, and predictive modeling to participate in the FluSight seasonal influenza forecasting challenge (Reich, McGowan, et al. 2019; Reich, Brooks, et al. 2019; Ray and Reich 2018; McGowan et al. 2019). Participants are encouraged to use historical surveillance data as well as weekly reports during the season to generate forecasts for influenza like illness (ILI) activity in the United States at the national and regional levels.

The dynamics of seasonal influenza and COVID-19 are undoubtedly distinct, with mitigation strategies and public health response to the spread of the diseases differing dramatically. Certain models that work well for seasonal flu may not be successful in terms of predicting the spread and severity of COVID-19. Nonetheless, many of the performers in the influenza modeling space have shifted focus to COVID-19. The pivot has been so significant that the 2019-2020 FluSight challenge was suspended prematurely in March 2020 and the 2020-2021 competition was not hosted. The Reich Lab at the University of Massachusetts-Amherst has been one of the leaders in developing collaborative infrastructure and novel analysis methods used in the FluSight effort. Much of this infrastructure and methodology has been re-purposed in service of the COVID-19 Forecast

Hub consortium. The initiative, which is supported by the CDC and NIH, invites research teams to openly submit forecasts of COVID-19 in the United States. The required format for forecasts includes point estimates and binned distributions for endpoints including the cumulative number (and/or increase) in COVID-19 cases, deaths, and hospitalizations across counties, states, and nationally. Eligible near-term forecasts (1 to 4 week horizons) are combined in a weekly ensemble model, the results of which are reported by the CDC.^2^

Here we describe the **Fo**recasting **CO**VID-19 in the **U**nited **S**tates (**FOCUS**) project, an endeavor to develop an automated modeling implementation to generate forecasts for weekly submission to the COVID-19 Forecast Hub. The work completed under this effort includes implementation of an autoregressive forecasting approach, development of an open-source software tool and cloud-based automation pipeline, and evaluation of model performance. By the end of the project, the FOCUS team submitted more than three months of weekly forecasts for national and state level incident case, incident death, and cumulative death targets, all of which were included in the COVID-19 Forecast Hub ensemble modeling.

## 2 Methods

### 2.1 Data Sources

The first step for the FOCUS effort was to identify a reliable data source for incident and cumulative counts of cases and deaths in the United States. For our forecasts, we relied on daily count data curated in the COVID-19 Data Repository by the Center for Systems Science and Engineering (CSSE) at Johns Hopkins University (Dong, Du, and Gardner 2020). In initial efforts, we also explored the New York Times^3^ COVID-19 repository, however we chose to solely use the JHU CSSE data given that the COVID-19 Forecast Hub organizers consider it the source of truth data for model evaluations.

During the FOCUS project period, the JHU CSSE group maintained daily count data posted as `.csv` files in a GitHub repository.^4^ Data were collated from public health agencies across the United States, and were made available at the county level. We developed code to programmatically read the `.csv` files with confirmed cases and deaths directly from GitHub, and then aggregate daily data to weekly counts. The code is written to parameterize granularity at which the counts are aggregated. For example, a state-level aggregation would add up all daily cumulative counts from counties to arrive at a daily cumulative count for the state. From there, the cumulative count is translated to an incident count by subtracting the cumulative count of the prior day from that of the current day. Daily incident counts are summed to weekly incidence, from which weekly cumulative counts are then calculated. Note that each week is translated to “epiweek” (MMWR week)^5^ format prior to modeling.

### 2.2 Statistical models

We employed a time series approach (SigSci-TS) to forecast incident cases, incident deaths, and cumulative deaths. For case data, we first retrieved the JHU CSSE weekly incident case counts using the methods previously described. We then fit a non-seasonal autoregressive integrated moving average (ARIMA) model using the historical case data. Parameters for `p`, `d`, and `q` (i.e. the orders for autogreressive terms, differencing, and moving average respectively) were selected using an automated parameter tuning process implemented in the `fable` R package (O’Hara-Wild, Hyndman, and Wang 2020a). After automatically performing a grid search of possible values for each order using the Hyndman-Khandakar algorithm (Hyndman and Khandakar 2008), the ARIMA model will use the combination of parameters that minimizes the corrected Akaike information criteria (AICc) (Cavanaugh 1997). Given that the process is automated and driven by the weekly dynamics of the input data, the specific parameters chosen for the incident case ARIMA model may differ weekly and/or by location. It is also worth noting that the `fable` R package allows for the parameter space to be arbitrarily constrained. Our initial models of national targets only used an unconstrained parameter space. After we included state-level targets we observed implausible forecast results for some states and subsequently restricted the space as follows: `p` = 1 or 2 ; `d` = 0, 1, or 2; `q` = 0. The parameters for `P`, `D`, and `Q` (seasonal orders) were all set to 0 to achieve an ARIMA fit without seasonality. After fitting the ARIMA model(s) each week (one for each location being included in the submission), we generated weekly forecasts of incident cases out to a four week horizon.

For incident death forecasts we used a time series linear modeling framework (TSLM). While the TSLM model can account for time series components (e.g. trend or seasonality), we did not specify any exogenous regressors. The fit for incident deaths used a three-week lag of incident case counts as the only predictor. Forecasts out to the four week horizon were generated by passing incident case counts at each horizon. Given that the lag does span the fourth week, we had to sequence forecasting to first generate one week-ahead incident case forecasts in order to create incident death forecasts for all targets. Forecasts of cumulative deaths were generated by summing the cumulative number of incident deaths reported the prior week with each of the incident death forecasts through the four week horizon.

All forecasts yielded point estimates as well as predicted values that divide the distribution at the following quantiles: 0.01, 0.025, 0.05, 0.1, 0.15, 0.2, 0.25, 0.3, 0.35, 0.4, 0.45, 0.5, 0.55, 0.6, 0.65, 0.7, 0.75, 0.8, 0.85, 0.9, 0.95, 0.975, and 0.99. Quantile estimates were generated by first converting quantiles to the bounds of the associated prediction interval, then creating intervals of each desired width before finally joining the predicted limits back to the appropriate quantiles. The quantiles above were chosen to meet the COVID-19 Forecast Hub submission requirements.^6^ To ensure non-negative estimates, predicted values were truncated at 0.

We fit incident case and incident death models using the `ARIMA()` and `TSLM()` functions from the `fable` R package (O’Hara-Wild, Hyndman, and Wang 2020a). Forecasts from fitted models were generated using the `forecast()` function in the `fabletools` R package (O’Hara-Wild, Hyndman, and Wang 2020b). We arrived at prediction intervals by using the `hilo()` function originally released in the `distributional` R package (O’Hara-Wild and Hayes 2020) and re-exported in `fabletools`.

### 2.3 Automation

All code to retrieve, pre-process, analyze, and format submission data was written in R (R Core Team 2019) and packaged in `focustools`.^7^ The package provides documentation for all functions and is built to include internal data, such as the names, abbreviations, and FIPS codes for the state, territory, and county locations in the United States. Using the `focustools` functions in R, team members were able to interactively generate forecasts and prepare the submission format using the quantile format described previously.

All of the forecasting code for the project was maintained under version control. Prior to interactively generating forecasts, team members could make sure they had up-to-date versions of the code. However, the interactive forecast workflow could become redundant or problematic if multiple team members were to independently create the weekly forecast submission files. It is possible that despite a common interface, individual users might pass arguments, enforce assumptions, or otherwise script the workflow such that they would arrive at different forecast results. To guard against this, we sought to automate as much of the forecasting workflow as possible.

The FOCUS automation relied heavily on cloud computing via Amazon Web Services (AWS). We leveraged several AWS features, including:

- Elastic Compute Cloud (EC2)^8^ to provision a templated computing instance that can run the forecast pipeline, as well as a separate instance to host a forecast exploration app.
- Launch templates^9^ to codify the provisioning of the pipeline and app instances.
- Elastic IP^10^ service to allocate and associate a static IP to the app instance.
- Simple Storage Service (S3)^11^ for storing outputs from the forecast pipeline as well as software/scripts needed to provision the EC2 instance.
- Identity Access Management (IAM)^12^ roles to provide access between services (i.e. Lambda to EC2 and EC2 to S3).
- Lambda^13^ to deliver a serverless function to launch the EC2 forecast pipeline.
- CloudWatch^14^ to manage the execution of the Lambda function on a weekly schedule.

### 2.4 Evaluation

Methodology for evaluating COVID-19 forecasts is rapidly evolving. With a range of targets, horizons, and locations, there are numerous endpoints to evaluate. Furthermore, the type of forecasting error may have differential meaning depending on the context. For example, either over or under prediction may be more costly in terms of public health impact, and the effect of one or the other may need to be taken into account during evaluation. In some cases a point estimate may be more beneficial given ease of interpretation. Lastly, a simple yes/no describing whether or not the observed value fell within a predicted range may be of interest.

We evaluated the performance of our forecasts using three complementary metrics that collectively address each of the scenarios above:

- Weighted Interval Score (WIS): Proper scoring rule for predictive distributions that weights quantiles and penalizes overprediction and underprediction.
- Mean Absolute Error (MAE): Absolute difference between predicted point estimates and observed values.
- Interval coverage: Binary as to whether or not the observed count fell within the forecasted interval.

The WIS was developed by some of the coordinators of the COVID-19 Forecast Hub as a mechanism to evaluate submissions. Details of the methods to calculate the WIS have been described previously (Bracher et al. 2021) and contextualized with other evaluation mechanisms such as MAE and interval coverage (Cramer et al. 2021). All of these metrics depend on a reliable data source for observed counts. The most widely used source of truth for COVID-19 outcomes in the United States is the repository curated by the JHU CSSE group (Dong, Du, and Gardner 2020).

The WIS, MAE, and interval coverage metrics are all calculated for COVID-19 Forecast Hub submissions by the Carnegie Mellon University Delphi group.^15^ The scores are made available in a publicly-accessible AWS S3 bucket. We retrieved the pre-computed Delphi scores to evaluate the performance of our submitted forecasts. Scores were retrieved on 2021-05-10 and limited to submissions between January 11, 2021 and April 12, 2021. Each combination of forecast date, target, horizon, and location was assigned the following: WIS, MAE, and binary indicators as to whether or not the 50% and 95% prediction intervals covered the observed count. We summarized each metric by target, averaging across locations, forecast dates, and/or horizons. To rank the performance with respect to other participants, we computed a relative metric for WIS, MAE, and interval coverage by dividing our scores (and those of other participants) by the scores for the COVIDhub baseline model, which forecasts median incidence equal to the previous week (Cramer et al. 2021). For overall rankings (averaged across all locations, horizons, and forecast dates), we only included models that had at least 500 forecasts. The overwhelming majority of forecasts were submitted on Sundays or Mondays. All Sunday forecast dates were coerced to Monday in order to join with baseline model. A very small number of forecasts were not submitted on Sunday or Monday; the scores from these were not included in rankings. Code to retrieve and aggregate scores for rankings is available in the Appendix (A1).

## 3 Results

### 3.1 Forecasts submitted

Between January 11, 2021 and April 12, 2021 we issued 14 submissions to the COVID-19 Forecast Hub, totaling 6,660 forecasts across three targets (incident cases, incident deaths, and cumulative deaths), four horizons (1 to 4 weeks ahead), and 52 locations (national, 50 states, and Washington DC). Figure 1 shows the locations for which forecasts were submitted each week. While initial forecasts were only made on the national level, the majority of weekly submissions included forecasts for state-level targets as well. For some weeks, we excluded certain states prior to submission due to implausible estimates. The cause of these inconsistencies was typically a backlog in reporting of cases in the given state, and we did not feel that it was appropriate to submit a forecast based on unreliable data for that week. Every forecast submitted included estimates of values at required quantiles and point prediction. All submissions were accepted by the COVID-19 Forecast Hub and included in the weekly ensemble models reported by the CDC.

**Figure 1:**
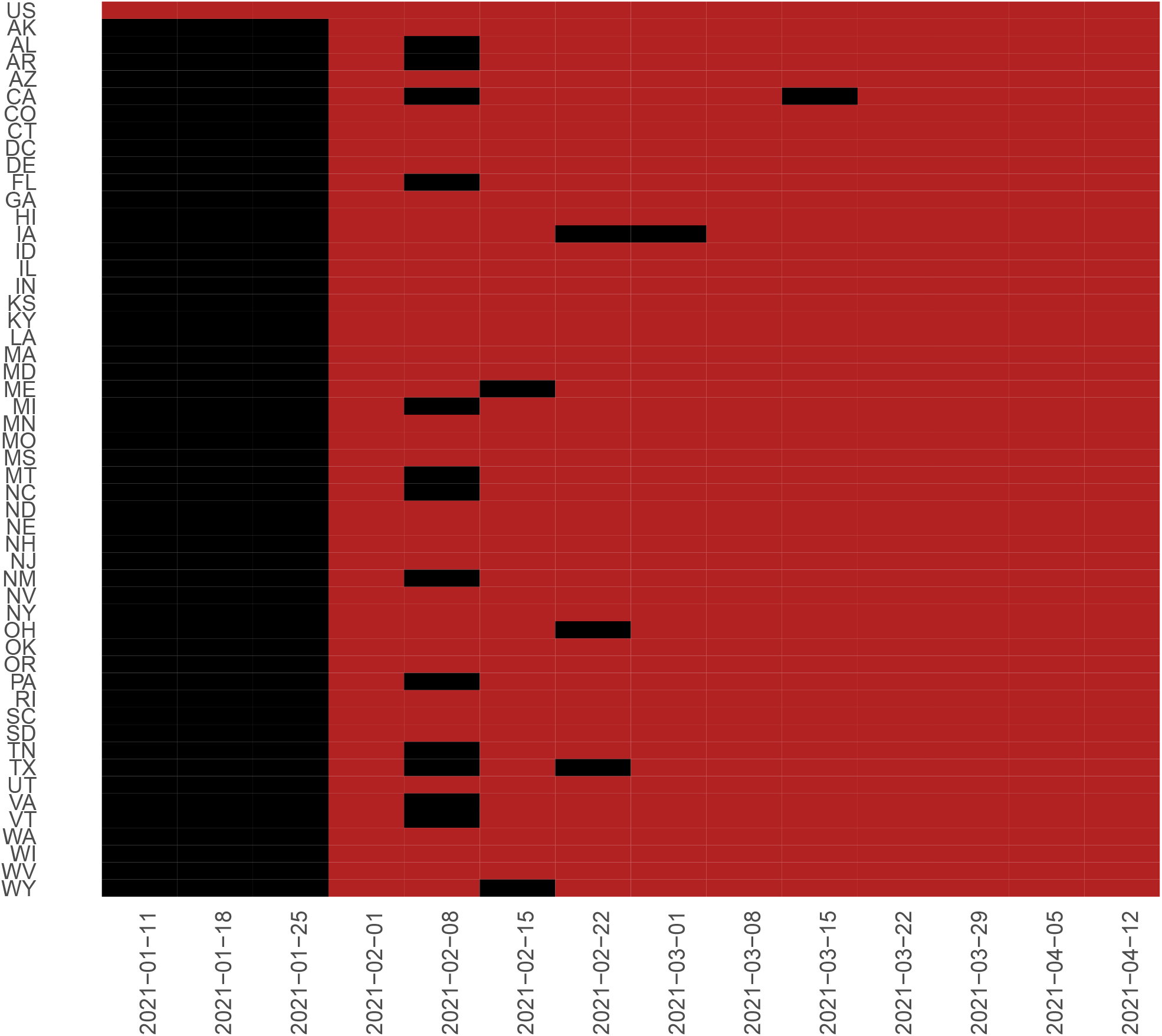
Weekly submission by location. Cells marked in red had 1-4 week horizons for incident case, incident death, and cumulative death targets included in the submission for the given week. Cells marked in black denote locations that were excluded from the weekly submission.

### 3.2 Automation

All functions to run statistical models, generate forecast outputs, prepare formatted submission files, and create tabular/visual summaries of forecasts are included in the `focutools` R package. The package is available on GitHub under the GPL-3 license and includes robust function documentation and narrative vignettes describing basic usage.^16^ By the end of the project, the package was used as part of the automated cloud pipeline. Figure 2 depicts the automation pipeline as implemented.

**Figure 2:**
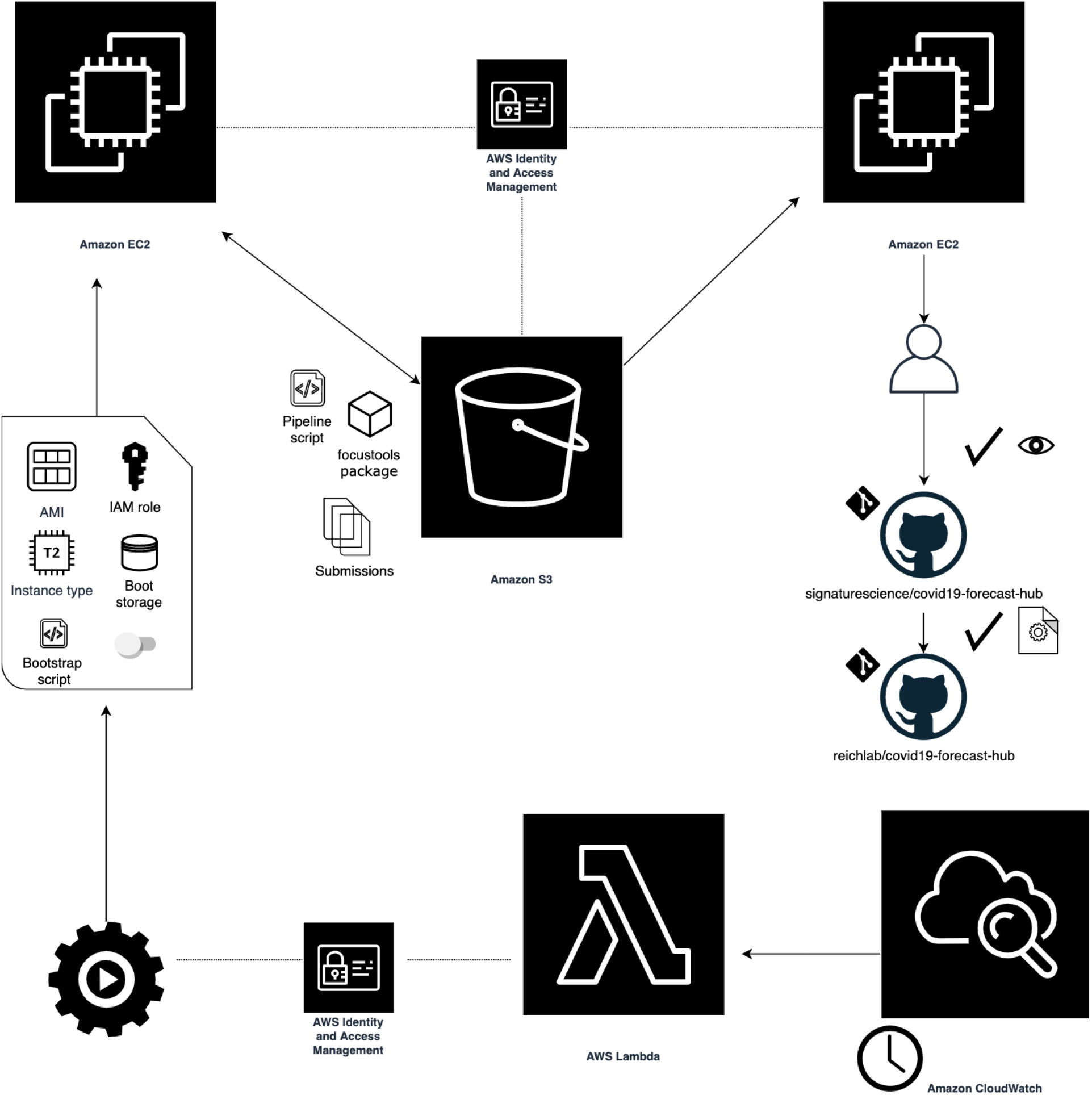
Illustration of automation pipeline. The weekly workflow starts with a scheduled AWS CloudWatch event to trigger an AWS Lambda operation. The function can access IAM credentials to launch the pipeline instance from a template that includes bootstrapping code, storage specifications, and details for the instance to be run. When the forecasting instance starts, it installs necessary software, generates forecast output, writes submission-ready files to an S3 bucket, and then self-terminates. Another EC2 instance hosts a web app with access to the same S3 bucket, allowing users to interactively review and download forecast output prior to submission. After reviewing through the web app, users manually inspect and then download the validated submission file, commit the file to the fork of the COVID-19 Forecast Hub repo, and submit a pull request upstream, which triggers an automated validation process.

The weekly forecasting workflow began with a CloudWatch event (defined by a cron rule) to trigger a Lambda function. The Lambda function used a Python script calling the `boto3`^17^ AWS client with the instruction to launch the pipeline instance from a template. The launch template included specific instructions needed to provision the instance: availability zone (us-east-1), instance type (T2.medium), AMI (Ubuntu 18.04 OS), IAM role with read/write access to the project S3 bucket, behavior on shutdown (terminate), and a definition of additional storage to be attached (16GB). The template also included a Base64 string that encoded a Bash bootstrapping script. Details of the steps performed in the bootstrap are available in the Appendix (A2).

The design for the weekly forecasting pipeline prioritized ephemerality. The bootstrap commands were run once, and by the end of the script the EC2 instance that ran the workflow was terminated. At the same time the following week, a new instance was launched with an identical configuration and set of instructions. Each weekly run used the same S3 bucket, from which team members could retrieve error logs if there were pipeline failures.

Once the forecasts were generated, the contents were reviewed by the team for plausibility of estimates. Team members reviewed plots of forecast output and tabular summaries of the predictions. To streamline this review process, the automation stack included a second instance to host a web interface for visually inspecting, filtering, and downloading forecast submission files. The web interface was written in R as a Shiny application and delivered via a wrapper function in the `focustools` R package. As with the forecast pipeline, details of the instance hosting the app were defined in a launch template: availability zone (us-east-1), instance type (T2.small), AMI (Ubuntu 18.04 OS), IAM role with read/write access to the project S3 bucket, and a definition of additional storage to be attached (16GB). Details of the bootstrapping steps performed on instance launch are available in the Appendix (A3).

Note that unlike the pipeline instance, the bootstrap commands for the app host do not end with a terminated instance. In fact, the instance hosting the app was assigned a static IP allocated via the Elastic IP service and stayed running such that team members could access the interface at their convenience. The machine was configured to run a cron job every hour to refresh the contents of a directory synced from S3 containing all pipeline-generated forecast files. The app listed the contents of that directory and presented each file name as a selection in a drop-down. After selecting a forecast file from the drop-down, the user could view plots of the forecasted curve and review tables of predicted counts and percent change over the four week horizon. Each result was faceted to locations selected by the user (defaulting to all locations present in forecast file). When ready to submit, the user could export the validated `.csv` submission file directly from the web interface. Figure 3 is a screenshot of the FOCUS explorer app interface.

**Figure 3:**
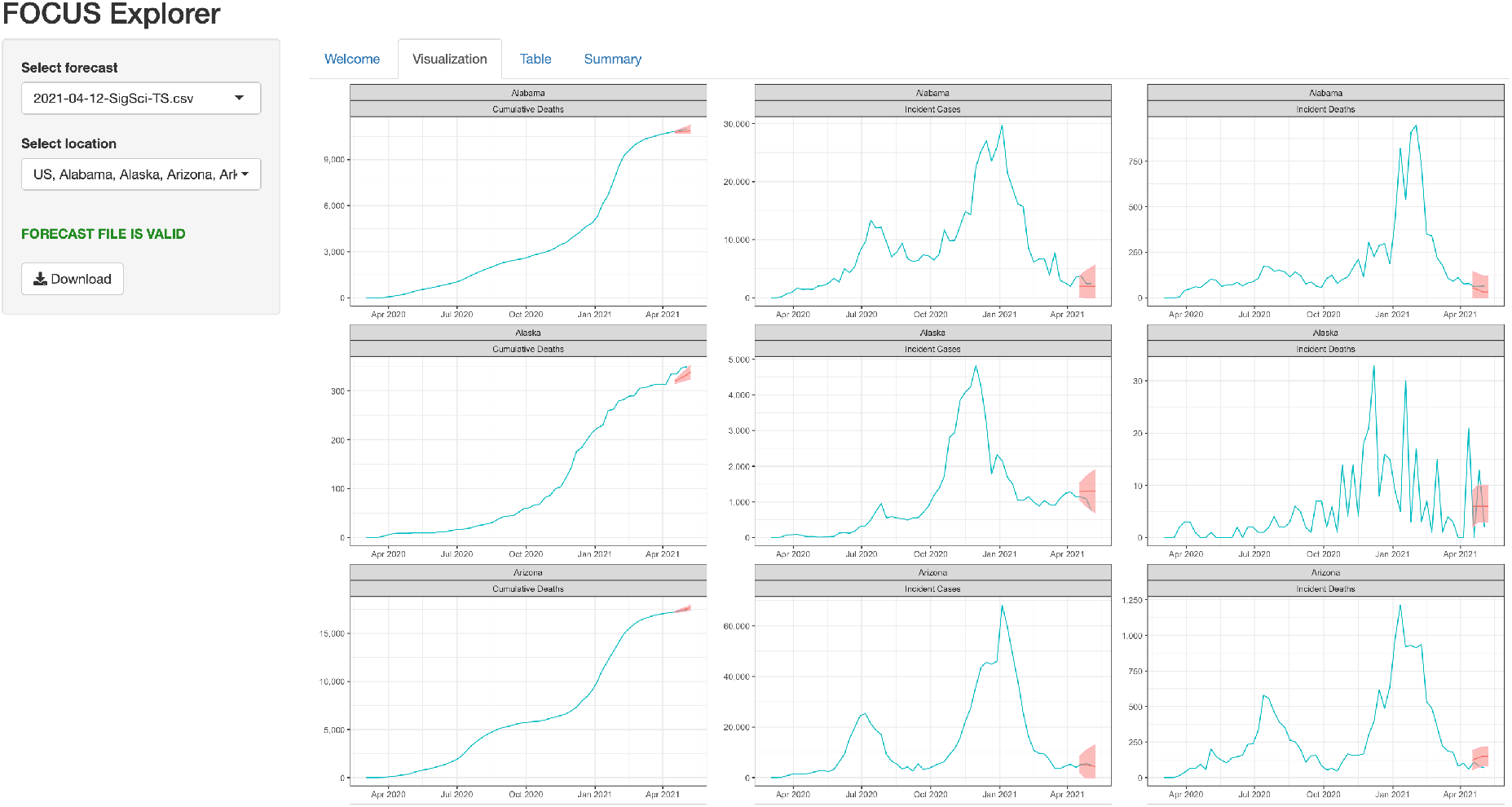
Screenshot of FOCUS Explorer app. The FOCUS Explorer Shiny application ships with the focustools R package and can be run locally or can be hosted. When starting the Shiny application, users must provide historic time series data prepared with the data retrieval functions from the focustools package. The app allows users to interactively explore forecast output through visualizations of observed counts and forecasts and tabular summaries. Each plot is stratified by location and target. The user can restrict locations and download the data in validated COVID-19 Forecast Hub submission format.

### 3.3 Scoring

Table 1 details how SigSci-TS forecasts submitted between 2021-01-11 and 2021-04-12 ranked relative to other submissions to the COVID-19 Forecast Hub. Metrics were averaged across all locations (US and states/territories) and horizons (1-4 weeks). Models were only ranked if they had scores available for at least 500 forecasts (combined across locations, horizons, and forecast dates). In terms of WIS, our forecasts ranked 6 of 30 for incident cases and 16 of 41 for incident death targets. When evaluating with MAE on average the forecasts ranked 3 of 33 for incident cases and 39 of 45 for incident death targets. For coverage, 95.2% and 75% incident case forecasts included the observed count in the 95% and 50% prediction intervals respectively. 93.1% and 48.5% incident death forecasts included the observed count in the 95% and 50% prediction intervals respectively. Our incident case model ranked 3 of 29 in the 95% coverage and 1 of 29 in the 50% coverage. The incident death forecasts ranked 3 of 42 in the 95% coverage and 9 of 42 in the 50% coverage.

**Table 1:** Rankings for SigSci-TS forecasts based on scores averaged across all forecast dates (2021-01-11 through 2021-04-12), locations, and horizons. Teams with less than 500 forecasts were excluded from rankings.

| Score | Incident Cases | Incident Deaths |
| --- | --- | --- |
| Coverage (50%) | 1 of 29 | 9 of 42 |
| Coverage (95%) | 3 of 29 | 3 of 42 |
| MAE | 3 of 33 | 39 of 45 |
| WIS | 6 of 30 | 16 of 41 |

The rankings above were computed by averaging across all horizons. Figures 4-7 illustrate SigSci-TS performance in incident death and incident case forecasts for WIS and MAE metrics stratified by horizon (1-4 weeks).

**Figure 4:**
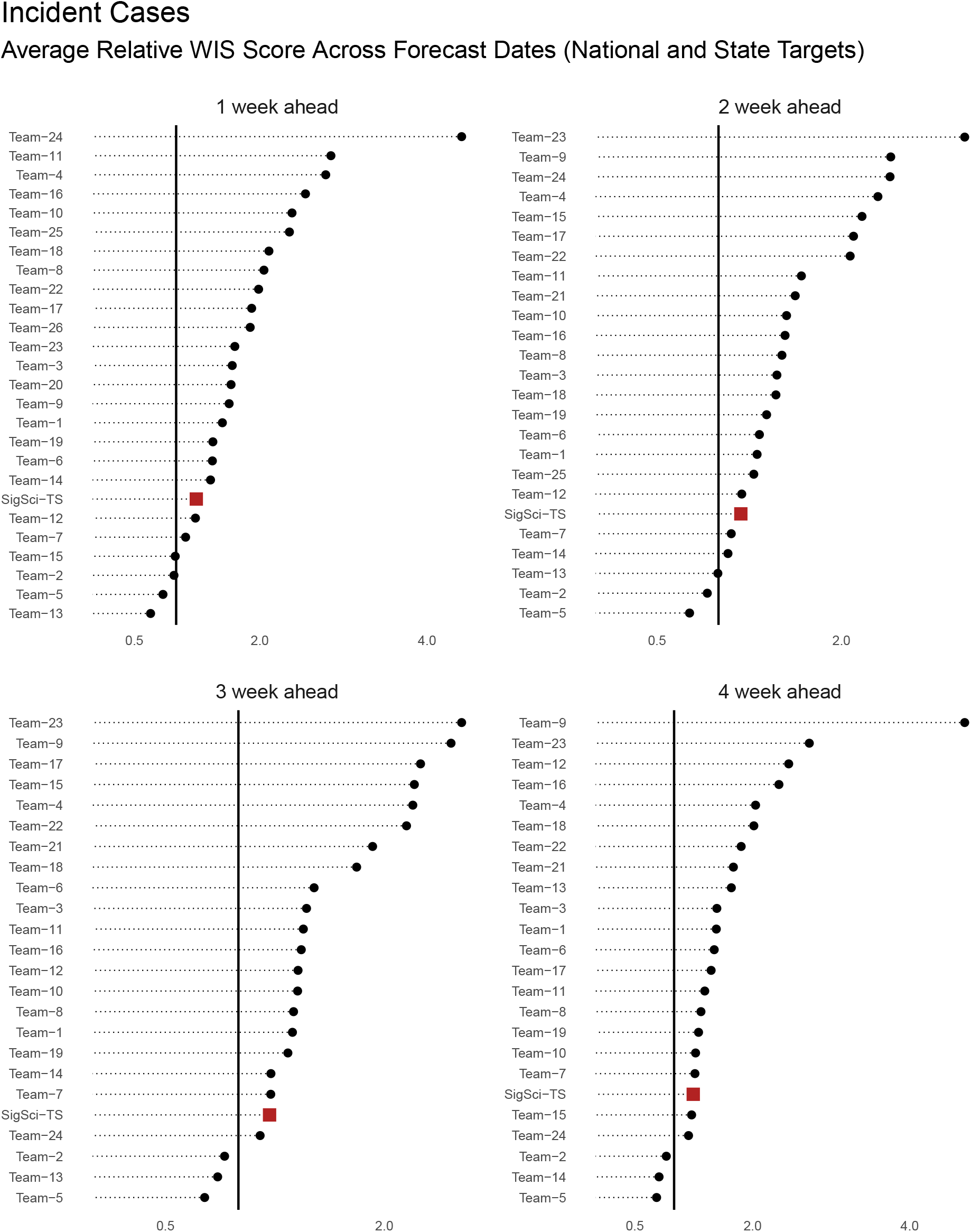
WIS rankings for incident case forecasts stratified by horizon. Scores are averaged across all locations and forecast dates (between 2021-01-11 and 2021-04-12) and calculated relative to the COVIDhub-baseline model. For this plot teams with less than 125 forecasts were excluded. One team was excluded from the plot because of extreme outlying scores. SigSci-TS is represented as a red square. The horizontal black line represents marks the relative score of 1 (i.e. the baseline model performance).

**Figure 5:**
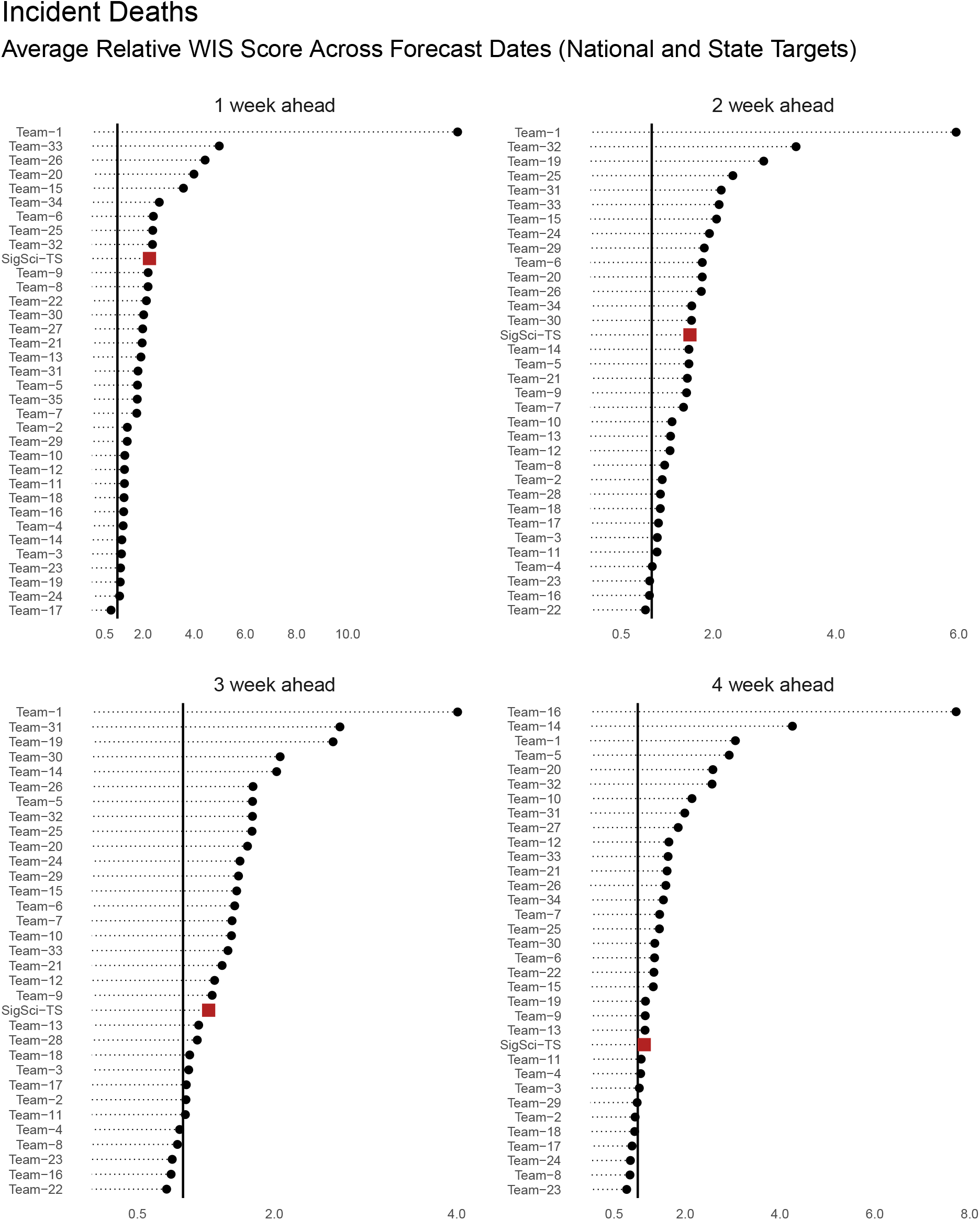
WIS rankings for incident death forecasts stratified by horizon. Scores are averaged across all locations and forecast dates (between 2021-01-11 and 2021-04-12) and calculated relative to the COVIDhub-baseline model. For this plot teams with less than 125 forecasts were excluded. One team was excluded from the plot because of extreme outlying scores. SigSci-TS is represented as a red square. The horizontal black line represents a relative score of 1 (i.e. the baseline model performance).

**Figure 6:**
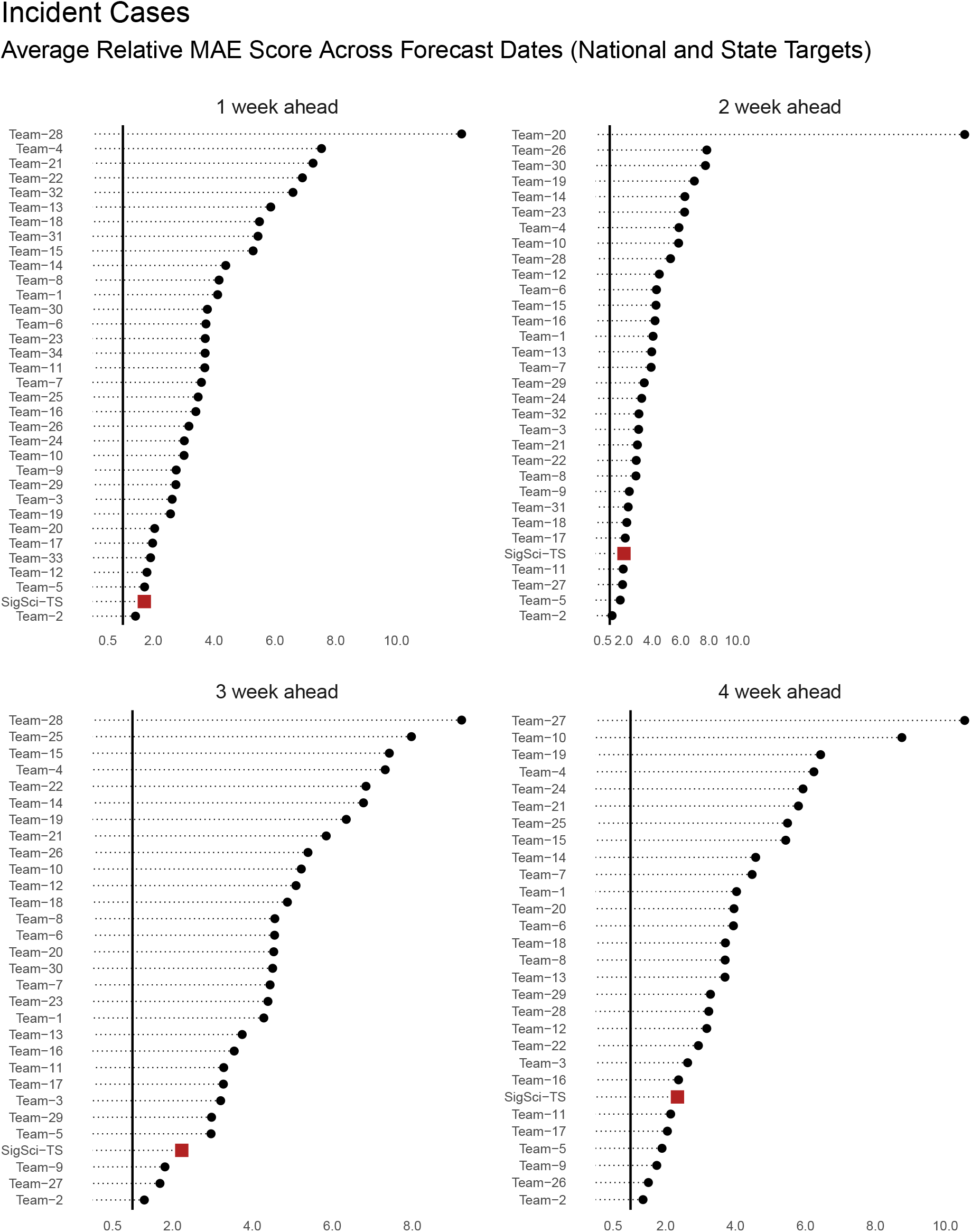
MAE rankings for incident case forecasts stratified by horizon. Scores are averaged across all locations and forecast dates (between 2021-01-11 and 2021-04-12) and calculated relative to the COVIDhub-baseline model. For this plot teams with less than 125 forecasts were excluded. Two teams were excluded from the plot because of extreme outlying scores. SigSci-TS is represented as a red square. The horizontal black line represents a relative score of 1 (i.e. the baseline model performance).

**Figure 7:**
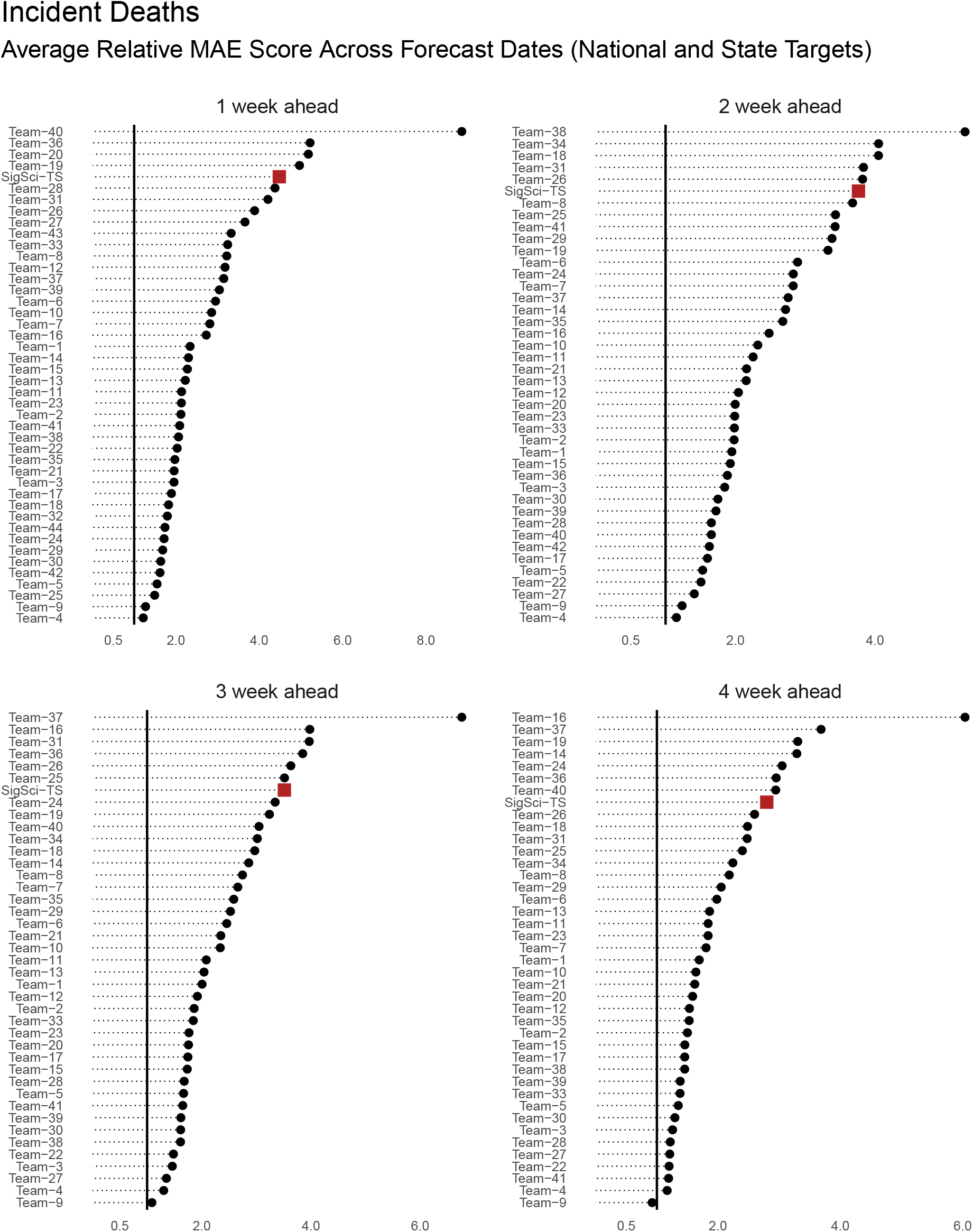
MAE rankings for incident death forecasts stratified by horizon. Scores are averaged across all locations and forecast dates (between 2021-01-11 and 2021-04-12) and calculated relative to the COVIDhub-baseline model. For this plot teams with less than 125 forecasts were excluded. Two teams were excluded from the plot because of extreme outlying scores. SigSci-TS is represented as a red square. The horizontal black line represents a relative score of 1 (i.e. the baseline model performance).

## 4 Discussion

The FOCUS project successfully forecast COVID-19 targets across a range of horizons and locations in the United States. All weekly forecasts submitted were included in ensemble models of targets at the national and state levels. We feel confident that the forecasts submitted contributed useful information to the ensemble, and were generally plausible and meaningful on their own. When summarized across all locations and horizons, the SigSci-TS approach ranked well above average in almost all of the evaluation metrics. The incident case and incident death models performed particularly well in the percent coverage metrics. While the coverage metric does not summarize forecast precision, it is useful for public health messaging. It is worth noting that the forecasts that “cast a wide net” (i.e. generate extremely broad prediction intervals) could perform well in this metric without being truly meaningful. For example, a forecast with a 95% prediction interval of between 0 and 10,000,000 weekly cases in the United States would capture the true incidence (e.g. 500,000 cases) but the range would be implausible. However, one would expect that such forecasts would not demonstrate precision in point estimates via the MAE metric and would be penalized for overprediction or underprediction in WIS scoring. The SigSci-TS ranked among the best for incident case models, performing well on percent coverage metrics and relatively well by the WIS and MAE metrics too.

One of the key features of the project was the automation of weekly forecasting. The modeling strategy that we developed has been fully implemented in the `focustools` R package such that our group, other users, and even an automated pipeline could generate formatted forecast outputs. The scheduled AWS pipeline saved time for the team members and ensured consistent structure and scheduling for forecast preparation. It is worth emphasizing that the pipeline was not fully automated through submission. Team member review of automatically generated forecast output via the explorer app was a critical step to ensure plausibility of estimates prior to submitting.

The FOCUS project had notable limitations. The SigSci-TS models used time series approaches. Autoregressive models have been shown to be successful in flu forecasting, and forecasts from our time series models generally appeared to score well for the period evaluated. However, we acknowledge that at different points in the pandemic (e.g. early on when there is very little data collected) a time series model may not be sufficient. Furthermore, while the SigSci-TS forecasts performed well for the near-term (1 to 4 week) horizons, we cannot and should not extrapolate performance evaluation to longer horizons or scenario projections. If applied to longer horizons, the SigSci-TS approach would have no way to bound predictions at the size of the susceptible population. While we only developed forecast submissions using time series methods, we did cursorily explore other approaches including compartmental models. Future work may involve development of a composite modeling approach calibrated to horizon. Such a framework might weight a time series model differently than another method (e.g. a compartmental model) given the number of weeks ahead for the forecast. In theory this could leverage the strengths of each constituent approach while mitigating model-specific limitations.

Beyond limitations specific to the SigSci-TS approach, there are general challenges to forecasting COVID-19 outcomes in the United States that are worth noting. In particular, data reporting standards have not been consistent across the United States during the pandemic. The JHU CSSE group has engaged in a massive undertaking to curate disparate data sources. In the process of doing so, the maintainers of the JHU CSSE repository have issued thousands of updates and errata to reported counts.^18^ From week to week, some state and county-level dashboards become “stale” and are not updated at expected intervals. Other locations have systematically shifted criteria for how cases and deaths are reported. As a result, in some cases training data may be a moving target and inconsistencies on any given week could influence weekly forecasts as well as evaluation at a later date.

## 5 Conclusion

The FOCUS project led to development of weekly forecasts of COVID-19 targets in the United States. All forecasts were submitted to the COVID-19 Forecast Hub, used in ensemble modeling, and reported by the CDC. The forecasting pipeline was developed as a software tool that was used as part of an automated pipeline.

## Data Availability

Code and data sources used in project execution and evaluation are linked as footnotes and references throughout the manuscript.

https://github.com/signaturescience/focustools

## 8. Appendix

### 8.1 A1: Code use to retrieve scoring data

The R script below is used to retrieve the scoring data from the Delphi Group’s Forecast Evaluation Dashboard (https://delphi.cmu.edu/forecast-eval/).

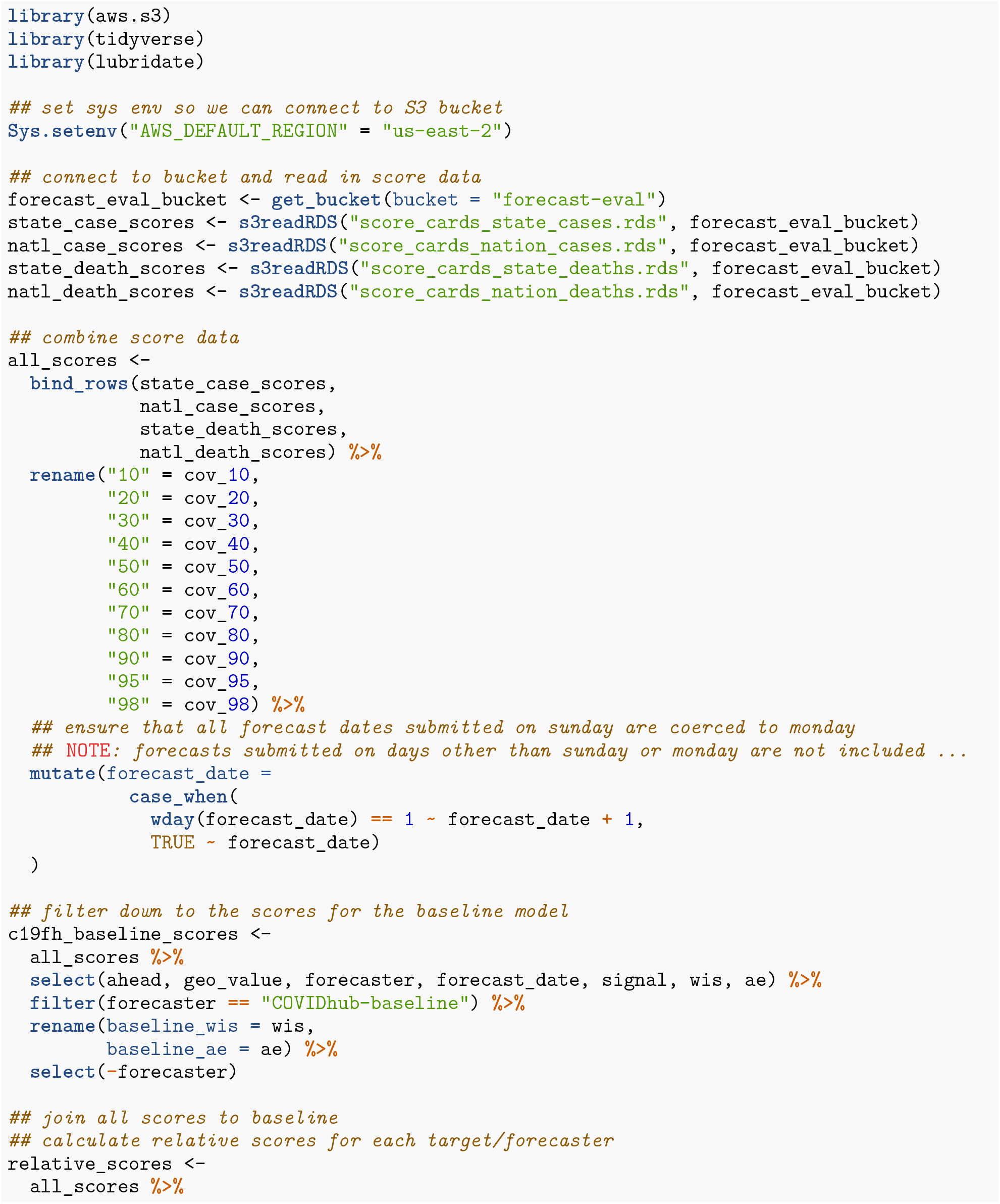
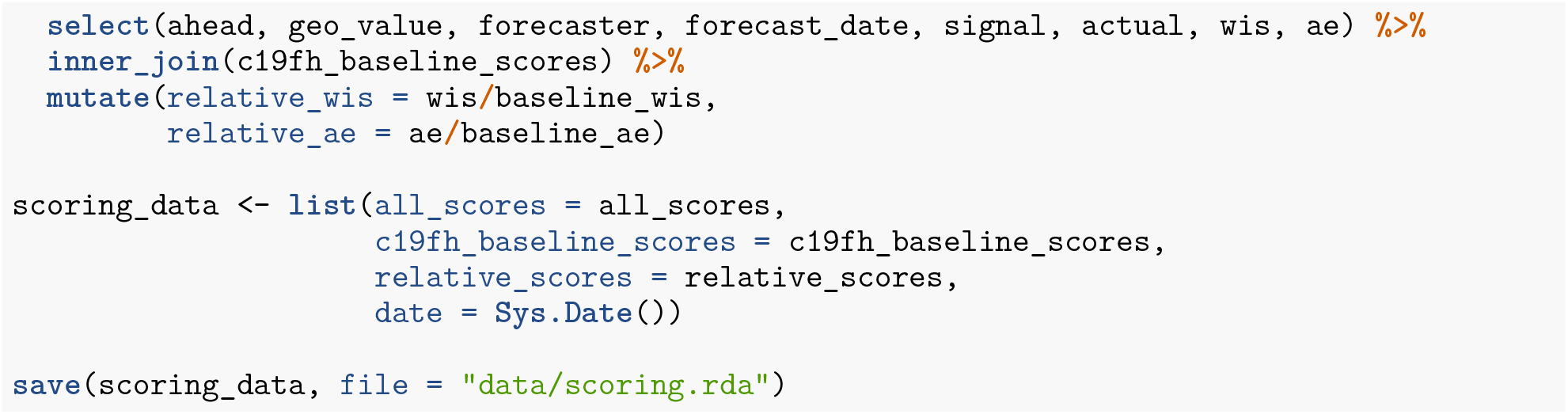

### 8.2 A2: Steps performed by bootstrap in the forecasting EC2 instance

1. Install R and system-level dependencies from the apt repository
2. Install the AWS CLI
3. Install Python dependencies for forecast validation
4. Install R package dependencies for the pipeline
5. Copy contents of the project S3 bucket to the host
6. Install the `focustools` R package from GitHub
7. Run the forecast pipeline R script (copied from S3)
8. Run validation check on output files
9. Copy forecast output files, validation results, and a log file to S3 bucket
10. Shut down the instance

### 8.3 A3: Steps performed by bootstrap in the app EC2 instance

1. Install R and system-level dependencies from the apt repository
2. Install the AWS CLI
3. Install Python dependencies for forecast validation
4. Install R package dependencies for the app
5. Copy contents of the project S3 bucket to the host
6. Install the `focustools` R package from GitHub
7. Configure cron for app refresh
8. Start the app

## 9. Acknowledgments

The work described in this manuscript built on collaborative efforts and public contributions from multiple entities. We acknowledge the following groups: JHU CSSE for daily COVID-19 data curation; the Reich Lab at the University of Massachusetts-Amherst for coordinating and developing the COVID-19 Forecast Hub infrastructure; the Delphi group at Carnegie Mellon University for calculating and distributing COVID-19 Forecast Hub performance scores; the CDC for guidance, interpretation, and dissemination of COVID-19 Forecast Hub forecasts; all participating teams in the COVID-19 Forecast Hub for contributions to the larger effort to understand the dynamics of the COVID-19 pandemic.

The FOCUS project was funded internally by Signature Science, LLC.

## Footnotes

2 https://www.cdc.gov/coronavirus/2019-ncov/covid-data/forecasting-us.html

3 https://github.com/nytimes/covid-19-data

4 https://github.com/CSSEGISandData/COVID-19/tree/master/csse_covid_19_data/csse_covid_19_time_series

5 https://www.n.cdc.gov/nndss/document/MMWR_week_overview.pdf

6 The COVID-19 Forecast Hub did not require that all targets have the full range of quantiles. Incident case forecasts required a subset: 0.025, 0.100, 0.250, 0.500, 0.750, 0.900, and 0.975. Additional details regarding COVID-19 Forecast Hub quantile forecast guidelines are available at https://github.com/reichlab/covid19-forecast-hub#forecast-files

7 https://github.com/signaturescience/focustools

8 https://aws.amazon.com/ec2/

9 https://docs.aws.amazon.com/AWSEC2/latest/UserGuide/ec2-launch-templates.html

10 https://docs.aws.amazon.com/AWSEC2/latest/UserGuide/elastic-ip-addresses-eip.html

11 https://aws.amazon.com/s3/

12 https://aws.amazon.com/iam/

13 https://aws.amazon.com/lambda/

14 https://aws.amazon.com/cloudwatch/

15 https://delphi.cmu.edu/forecast-eval/

16 https://signaturescience.github.io/focustools/

17 https://boto3.amazonaws.com/v1/documentation/api/latest/index.html

18 https://raw.githubusercontent.com/CSSEGISandData/COVID-19/master/csse_covid_19_data/csse_covid_19_time_series/Errata.csv

## Notes

### Competing Interest Statement

The authors have declared no competing interest.

### Funding Statement

The work presented in this manuscript was funded internally by Signature Science, LLC. No external funding was received.

### Author Declarations

The work described here did not involve human subjects and did not require IRB oversight/approval.

## References

Bracher, Johannes, Evan L. Ray, Tilmann Gneiting, and Nicholas G. Reich. 2021. “Evaluating Epidemic Forecasts in an Interval Format.” Edited by Virginia E. Pitzer. PLOS Computational Biology 17 (2): e1008618. https://doi.org/10.1371/journal.pcbi.1008618.

Cavanaugh, Joseph E. 1997. “Unifying the Derivations for the Akaike and Corrected Akaike Information Criteria.” Statistics & Probability Letters 33 (2): 201–8. https://doi.org/10.1016/S0167-7152(9600128-9).

Cramer, Estee Y., Evan L. Ray, Velma K. Lopez, Johannes Bracher, Andrea Brennen, Alvaro J. Castro Rivadeneira, Aaron Gerding, et al. 2021. “Evaluation of Individual and Ensemble Probabilistic Forecasts of COVID-19 Mortality in the US.” medRxiv, February, 2021.02.03.21250974. https://doi.org/10.1101/2021.02.03.21250974.

Dong, Ensheng, Hongru Du, and Lauren Gardner. 2020. “An Interactive Web-Based Dashboard to Track COVID-19 in Real Time.” The Lancet. Infectious Diseases 20 (5): 533–34. https://doi.org/10.1016/S1473-3099(20)30120-1.

Hyndman, Rob J., and Yeasmin Khandakar. 2008. “Automatic Time Series Forecasting: The Forecast Package for R.” Journal of Statistical Software 27 (3). https://doi.org/10.18637/jss.v027.i03.

McGowan, Craig J., Matthew Biggerstaff, Michael Johansson, Karyn M. Apfeldorf, Michal Ben-Nun, Logan Brooks, Matteo Convertino, et al. 2019. “Collaborative Efforts to Forecast Seasonal Influenza in the United States, 2015-2016.” Scientific Reports 9 (1): 683. https://doi.org/10.1038/s41598-018-36361-9.

O’Hara-Wild, Mitchell, and Alex Hayes. 2020. Distributional: Vectorised Probability Distributions. https://CRAN.R-project.org/package=distributional.

O’Hara-Wild, Mitchell, Rob Hyndman, and Earo Wang. 2020a. Fable: Forecasting Models for Tidy Time Series. https://CRAN.R-project.org/package=fable.

O’Hara-Wild, Mitchell, Rob Hyndman, and Earo Wang. 2020b. Fabletools: Core Tools for Packages in the ’Fable’ Framework. https://CRAN.R-project. org/package=fabletools.

Ray, Evan L., and Nicholas G. Reich. 2018. “Prediction of Infectious Disease Epidemics via Weighted Density Ensembles.” Edited by Cecile Viboud. PLOS Computational Biology 14 (2): e1005910. https://doi.org/10.1371/journal.pcbi.1005910.

R Core Team. 2019. R: A Language and Environment for Statistical Computing. Vienna, Austria: R Foundation for Statistical Computing. https://www.R-project.org/.

Reich, Nicholas G., Logan C. Brooks, Spencer J. Fox, Sasikiran Kandula, Craig J. McGowan, Evan Moore, Dave Osthus, et al. 2019. “A Collaborative Multiyear, Multimodel Assessment of Seasonal Influenza Forecasting in the United States.” Proceedings of the National Academy of Sciences 116 (8): 3146–54. https://doi.org/10.1073/pnas.1812594116.

Reich, Nicholas G., Craig J. McGowan, Teresa K. Yamana, Abhinav Tushar, Evan L. Ray, Dave Osthus, Sasikiran Kandula, et al. 2019. “Accuracy of Real-Time Multi-Model Ensemble Forecasts for Seasonal Influenza in the U.s.” Edited by Virginia E. Pitzer. PLOS Computational Biology 15 (11): e1007486. https://doi.org/10.1371/journal.pcbi.1007486.

